# Meta-Analysis of Breast Cancer Risk for Individuals with PALB2 Pathogenic Variants

**DOI:** 10.1101/2023.05.31.23290791

**Authors:** Thanthirige Lakshika M. Ruberu, Danielle Braun, Giovanni Parmigiani, Swati Biswas

## Abstract

**Background:** Pathogenic variants in cancer susceptibility genes can now be tested efficiently and economically with the wide availability of multi-gene panel testing. This has resulted in an unprecedented rate of identifying individuals carrying pathogenic variants. These carriers need to be counselled about their future cancer risk conferred by the specific gene mutation. An important cancer susceptibility gene is PALB2. Several studies reported risk estimates for breast cancer (BC) associated with pathogenic variants in PALB2. Because of the variety of modalities (age specific risk, odds ratio, relative risk, and standardized incidence ratio) and effect sizes of these risk estimates, a meta-analysis of all of these estimates of BC risk is necessary to provide accurate counselling of patients with pathogenic variants in PALB2. The challenge, though, in combining these estimates is the heterogeneity of studies in terms of study design and risk measure.

**Methods:** We utilized a recently proposed novel Bayesian random-effects meta-analysis method that can synthesize and combine information from such heterogeneous studies. We applied this method to combine estimates from twelve different studies on BC risk for carriers of pathogenic PALB2 mutations, out of which two report age-specific penetrance, one reports relative risk, and nine report odds ratios.

**Results:** The estimated overall (meta-analysis based) risk of BC is 12.80% by age 50 (6.11% − 22.59%) and 48.47% by age 80 (36.05% − 61.74%).

**Conclusion:** Pathogenic mutations in PALB2 makes women more susceptible to BC. Our risk estimates can help clinically manage patients carrying pathogenic variants in PALB2.

## 1 Introduction

Multi-gene panel testing has made it possible to test many cancer-susceptible genes affordably and efficiently (Plichta et al., 2016). This has enabled identification of an increasingly large number of patients carrying pathogenic variants in various cancer genes. This includes both well-studied high-risk genes such as BRCA1 and BRCA2 and relatively less-studied moderate-risk genes. PALB2 is one such moderate-risk gene whose mutations are considered to be a predisposing factor for breast cancer (BC) (Rahman et al., 2007; Wesoła and Jeleń, 2017; Poumpouridou and Kroupis, 2012) as well as other cancers (Jones et al., 2009). PALB2 mutations have been identified in approximately 0.6–2% of familial BC (Hofstatter et al., 2011; Bogdanova et al., 2011; Dansonka-Mieszkowska et al., 2010).

Several studies have examined the impact of pathogenic variants in PALB2 on BC risk. Antoniou et al. (2014)conducted a family-based study of 364 female PALB2 carriers from 154 families to estimate the age-specific BC risk (penetrance) for PALB2 carriers. They found that the risk of BC for these carriers compared to a general population was higher by eight to nine times among those younger than 40 years of age, six to eight times among those 40 to 60 years of age, and five times among those older than 60 years of age. Further, they estimated that the cumulative risk of BC for female PALB2 carriers was 14% (CI; 9 - 20) by 50 years of age and 35% (CI; 26 - 46) by 70 years of age. Another study (Casadei et al., 2011) reported that the risk of BC for women with PALB2 mutations is 2.3-fold (95% CI: 1.5 – 4.2) by age 55 and 3.4-fold (95% CI: 2.4 – 5.9) by age 85 compared to their female relatives without the mutation. Aloraifi et al. (2015) conducted a meta-analysis of several case-control studies with cases being women diagnosed with early onset BC (less than 50 years) and reported a pooled odds ratio of 21.4. There are several other studies reporting BC risk conferred by PALB2 mutations (Zheng et al., 2018; Rahman et al., 2007; Erkko et al., 2008; Kurian et al., 2017; Díaz-Zabala et al., 2022; Dorling et al., 2021). Overall, there is a substantial variability in reported estimates.

For clinical purposes, it is necessary to combine risk information available in this vast literature to determine the age-specific risk of BC, that is, penetrance of BC due to pathogenic variants in PALB2. This would greatly benefit individuals carrying such variants. However, there is a wide variability among the published studies in terms of study design (e.g., family-based, case-control) and type of reported result (e.g., age-specific penetrance, odds ratio (OR), relative risk (RR), standardized incidence ratio (SIR)). Standard random-effects meta-analysis methods cannot be applied due to the differences in the type of reported summary results across studies. Thus, a key challenge in this task is synthesising all available information on risk while accounting for heterogeneity in study design across studies.

We have recently developed a novel meta-analysis approach based on a Bayesian hierarchical random-effects model to obtain penetrance estimates (Ruberu et al., 2024). It integrates results from studies reporting different types of risk measures while accounting for uncertainties associated with such synthesis. We have also used it to estimate the age-specific penetrance of female BC among carriers of pathogenic variants in the ATM gene. In particular, the method can combine studies reporting age-specific penetrance, OR, RR, and SIR. To the best of our knowledge, this is the only method currently available in the literature to combine different types of risk measures while maintaining satisfactory statistical properties, in particular, low mean square error and coverage probabilities of interval estimates being close to the nominal (expected) confidence level. The only other method equipped to combine different types of risk measures is a fixed-effects approach (Marabelli et al., 2016), however, its interval estimates can suffer from under-coverage (Ruberu et al., 2024) and it does not address uncertainty quantification as completely as in Ruberu et al. (2024). In this article, we applied this novel Bayesian hierarchical random-effects model to conduct the first meta-analysis of penetrance of BC for pathogenic variants in the PALB2 gene.

## 2 Methods

### 2.1 Selection of Studies

First, we identified potential studies to include in the meta-analysis using a semi-automated natural language processing–based procedure for abstract screening (Deng et al., 2019). We also included additional studies using a Pubmed search of the following keywords in the title/abstract of the articles: “((((mutation) OR (variant)) AND (PALB2)) AND (breast cancer)) AND (germline)” up to 10 May 2023. Our inclusion and exclusion criteria were the same as described in Ruberu et al. (2024). Specifically, we included (1) family-based segregation analyses or epidemiological studies reporting cancer risk information in terms of age-specific penetrance, RR, or SIR and (2) case-control studies comparing BC patients with healthy subjects and reporting either OR or sufficient data to estimate the OR and its 95% CI. We excluded casecontrol studies that ascertained subjects based on family history of BC but did not adjust for this ascertainment criterion in their estimation procedure as that can introduce bias in estimates (Kraft and Thomas, 2000). Moreover, we adjusted reported OR for a study to include only those mutations that are pathogenic as per ClinVar (Landrum et al., 2018), whenever that information was available. This resulted in inclusion of 12 studies in our meta-analysis, as summarized in Table 1. More details about these studies can be found in Supplementary Materials (Table S1).

**Table 1:** Summary of studies included in the meta-analysis of BC penetrance for PALB2 mutations.

| Index | Study | Cases | Study Design | Sample Size | Risk and CI<br>(Input to<br>meta-analysis) |
| --- | --- | --- | --- | --- | --- |
| 1 | Antoniou et al. (2014) | One family member with PALB2 BRCA1/2 negative | Family-based segregation analysis | 311 | Penetrance Curve |
| 2 | Erkko et al. (2008) | Family of PALB2 carrier with BC | Family-based segregation analysis | 213 | Penetrance Curve |
| 3 | Rahman et al. (2007) | Familial BC BRCA1/2 negative | Case-control/family-based segregation analysis | 2007 | RR = 2.3<br>(1.4 - 3.9) |
| 4 | Kurian et al. (2017) | Female patients who underwent panel testing | Case control study | 95561 | OR = 3.39<br>(2.79 - 4.12) |
| 5 | Momozawa et al. (2018) | Unselected BC | Case-control study | 18292 | OR = 9.00<br>(3.4-29.7) |
| 6 | Díaz-Zabala et al. (2022) | Unselected BC | Case-control study | 3286 | OR = 3.76<br>(0.66 - 21.53) |
| 7 | Cybulski et al. (2015) | Recruited from hospitals with a subset of early onset BC (<50 years) | Case-control study | 17231 | OR = 4.39<br>(2.30-8.37) |
| 8 | Dorling et al. (2021) | Mainly unselected BC with a subset of early onset BC BRCA1/2 negative | Case-control study | 97997 | OR = 5.02<sup>a</sup><br>(3.73-6.76) |
| 9 | Heikkinen et al. (2009) | Sporadic BC | Case-control study | 2353 | OR = 3.4<sup>d</sup><br>(0.68-32.95) |
| 10 | Ahearn et al. (2022) | Recommended for biopsy or documented BC | Case-control study | 2434 | OR = 17.25<br>(2.15-138.13) |
| 11 | Zheng et al. (2018) | Unselected BC | Case-control study | 2133 | OR = 20.384<sup>b,c</sup><br>(NA) |
| 12 | Felix et al. (2022) | Familial BC | Case-control study | 290 | OR = 4.96<sup>b,c</sup><br>(NA) |

<sup>a</sup> Based on 30 studies in breast cancer association consortium (BCAC) unselected for family history.
<sup>b</sup> No mutations in controls
<sup>c</sup> OR and CI were calculated using data reported in the paper
<sup>d</sup> Only sporadic cases were considered

### 2.2 Statistical Methods

To perform the meta-analysis, we employed a novel approach based on a Bayesian hierarchical random-effects model as described in Ruberu et al. (2024). Briefly, we combine estimates from *S* studies reporting point estimates as well as standard errors in one of four modalities: penetrance estimates/curve in an age range (e.g. 35−85) or at few years such as 40, 50,…, 80, RR, SIR or OR. We assume that for all studies, the underlying cumulative risk *F*_*s*_(*t* |*κ*_*s*_, *λ*_*s*_) at age *t* is given by a Weibull distribution with study-specific parameters *κ*_*s*_ and *λ*_*s*_. Each type of risk measure is assumed to follow a normal distribution whose mean is a function of penetrance (i.e., Weibull parameters *κ*_*s*_ and *λ*_*s*_) and variance reported by the study.

The prior distributions used in our model were chosen to produce a wide and realistic range of age-specific risks, so to be applicable to most gene-cancer applications. Specifically, we used the following hierarchical priors: *κ*_*s*_ ∼ Gamma(*a, b*), *λ*_*s*_ ∼ Gamma(*c, d*), where *a* and *c* are shape parameters, *b* and *d* are scale parameters, and *a* ∼ U(7.5, 27.5), *b* ∼ U(0.15, 0.25), *c* ∼ U(43, 63), and *d* ∼ U(1.32, 2.02). The posterior distributions were estimated by a Markov chain

Monte Carlo algorithm, and used to estimate the consensus age-specific penetrance curve and its 95% credible interval. Additional details about the method are provided in the Supplementary Materials. We carried out all analyses in the statistical software system R (R Core Team, 2020). The method is implemented in the R package BayesMetaPenetrance, available at https://personal.utdallas.edu/~sxb125731/ and https://github.com/LakshikaRuberu.

## 3 Results

The penetrance estimate of BC for carriers of PALB2 pathogenic variants obtained by Bayesian meta-analysis is shown in Figure 1. The risk of BC is 12.80% by age 50 (6.11% − 22.59%) and 48.47% by age 80 (36.05% − 61.74%). For comparison, the figure also shows the age-specific risk estimates for non-carriers obtained using SEER (Surveillance, Epidemiology, and End Results) data. We also compared the meta-analysis curve with the estimates provided by each of the twelve individual studies in Figure 2, wherein for each study, we plot a Weibull penetrance curve that approximates the risk estimate provided by each study listed in Table 1. Note that the meta-analysis penetrance curve lies towards the center of the twelve curves. Notably, the contribution of each study to the final meta-analysis curve depends not only on the reported risk estimates but also on their standard errors (or confidence intervals) and sample sizes.

**Figure 1:**
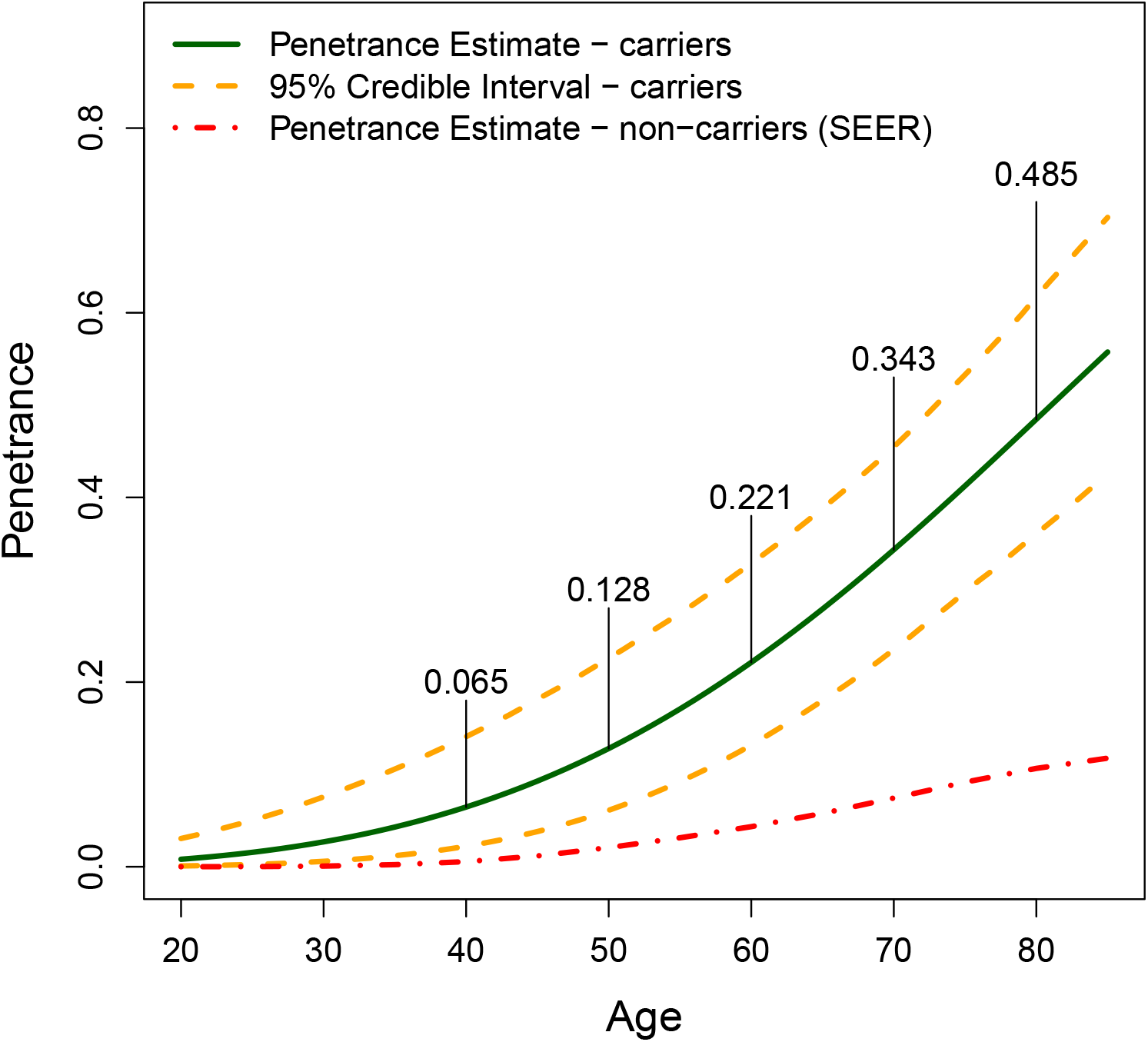
Penetrance estimate of BC for PALB2 mutations.

**Figure 2:**
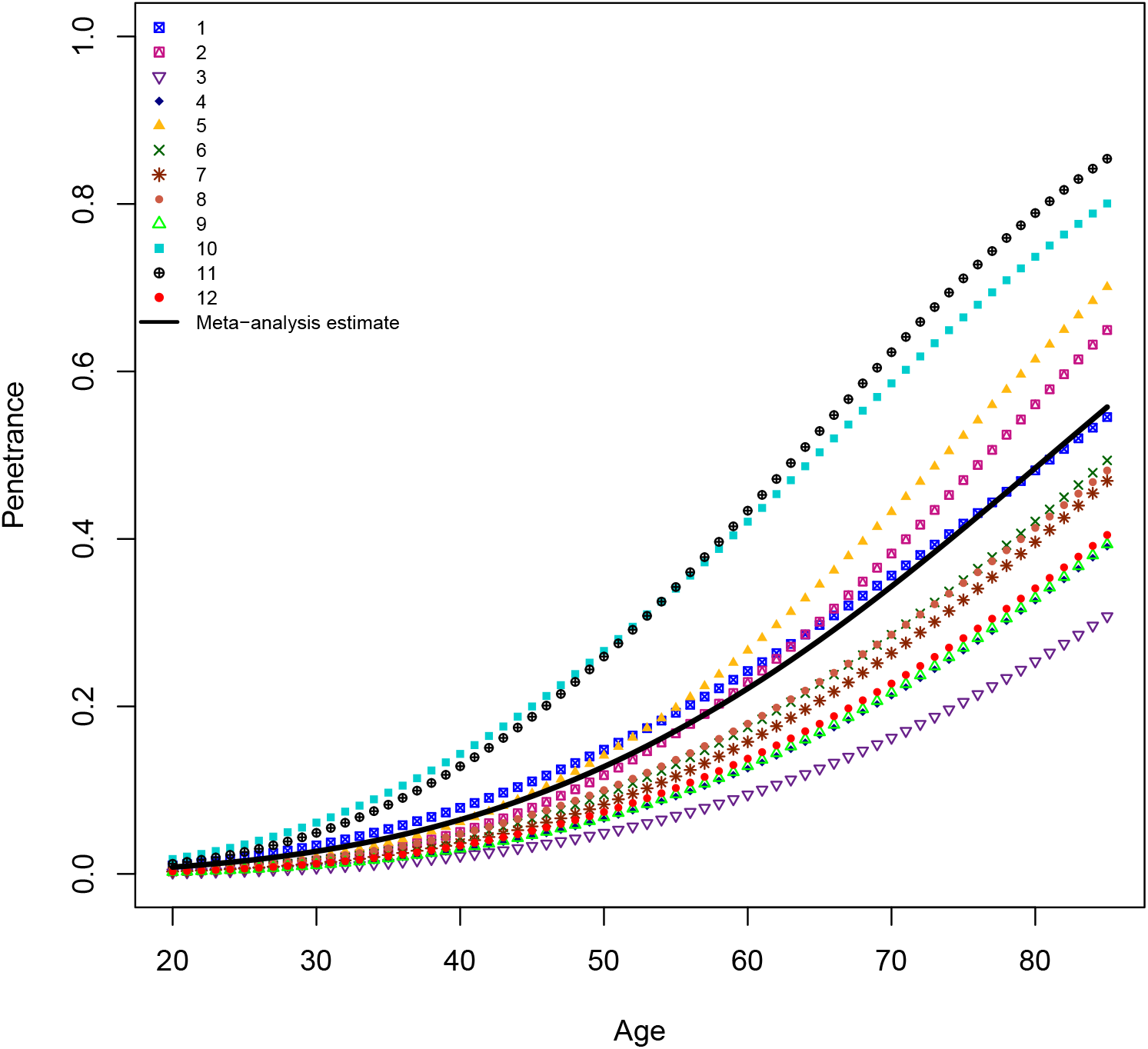
Approximate penetrance curves (Weibull distributions) for each of the 12 studies: The number in the legend corresponds to the study index in Table 1.

Among our 12 selected studies, two that reported OR included a subset of early-onset BC cases (Cybulski et al., 2015; Dorling et al., 2021) while one study included controls with ages greater than 60 (Momozawa et al., 2018). We investigated how sensitive our results are to these studies by removing them one at a time. The resulting three penetrance curves are very close to the curve based on all 12 studies in Figure 3(a) showing that our final curve is robust to the exclusion of these studies. We note that OR for two studies, Ahearn et al. (2022) and Zheng et al. (2018) are quite high (17.25 and 20.384, respectively). These were the only two studies from Western Africa, specifically, Ghana and Nigeria. To investigate how sensitive the overall penetrances are to these two studies, we repeated the meta-analysis after removing these studies one at a time. The resulting penetrance curves are shown in Figure 3(b). We can see that they are close to the overall penetrance curve.

**Figure 3:**
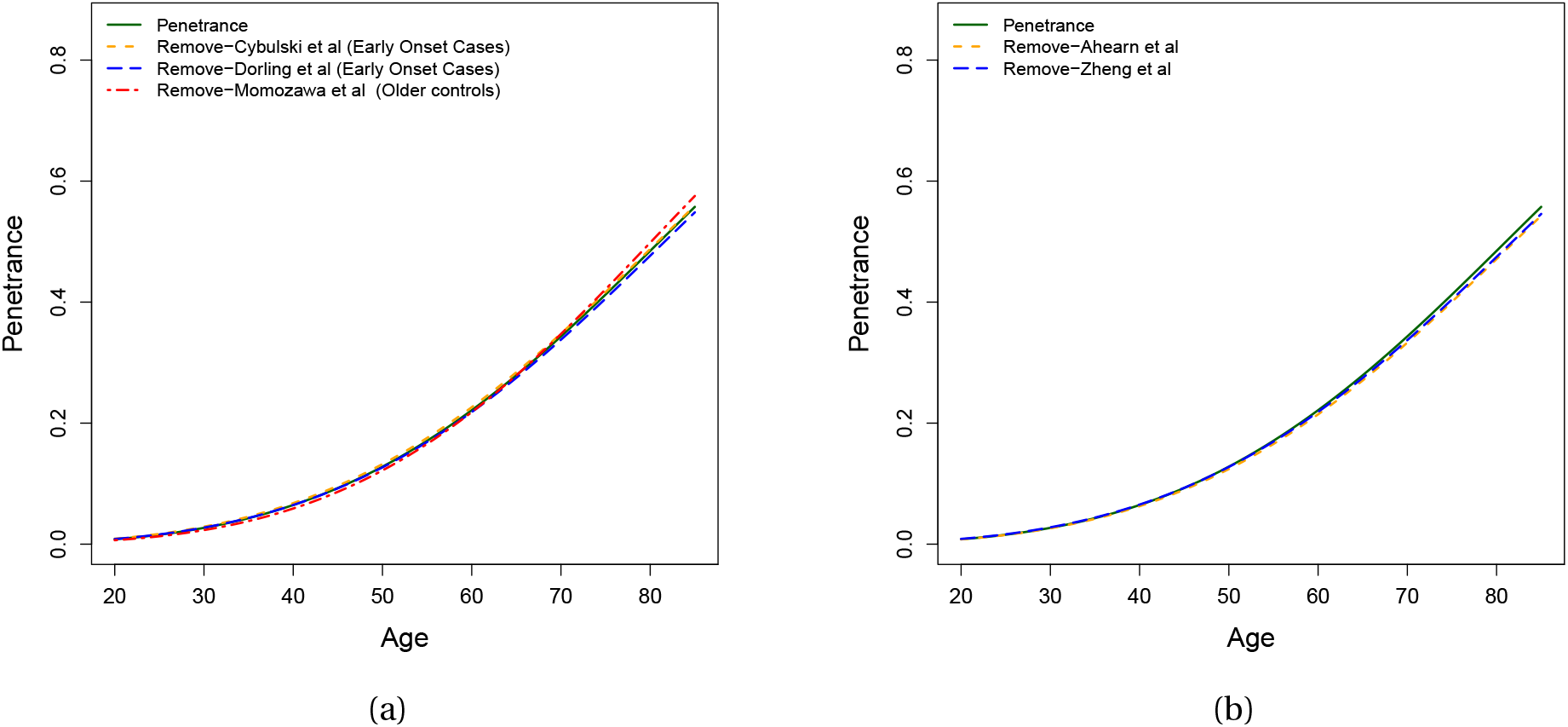
Results of sensitivity analysis: (a) Removing studies with early onset cases or controls restricted to older ages, one at a time. (b) Removing Ahearn et al. (2022) and Zheng et al. (2018) one at a time.

## 4 Discussion

Multi-gene panel testing has created an opportunity to identify people carrying mutations in a wide variety of cancer susceptibility genes (Plichta et al., 2016). PALB2 is one such gene which is now included in all BC gene-testing panels enabling identification of more people carrying pathogenic variants in the PALB2 gene (Tung et al., 2015). To counsel these carriers and help them mitigate their cancer risks, clinicians need estimates of age-specific risk (penetrance) associated with the pathogenic variants in PALB2. In this paper, we estimated penetrance for female BC by applying a novel Bayesian hierarchical random-effects meta-analysis method (Ruberu et al., 2024). To the best of our knowledge, this is the first meta-analysis to assess the penetrance of BC among female carriers of pathogenic mutations in PALB2.

Our results are consistent with the age-specific penetrance estimates for PALB2 mutations reported in the literature. Only two studies report age-specific penetrance estimates, namely Antoniou et al. (2014) and Erkko et al. (2008); both are included in our meta-analysis. As shown in Figure 2, the estimated meta-analytic penetrance curve lies between the two penetrance curves obtained from these studies.

We excluded studies that did not appropriately adjust for their ascertainment criteria based on family history in their analysis. Although we did include studies where participants were ascertained by age, our sensitivity analysis shows that the final estimates are robust to the inclusion of such studies. Also, we note that none of the two studies reporting penetrance provided results for ages less than 35. Moreover, none of the included OR/RR studies are age-specific. Therefore, the reported meta-analysis penetrance curve should be interpreted with caution for ages less than 35 due to the lack of direct empirical information.

A limitation of our meta-analysis is that we had to combine risk estimates from studies with different ancestries/populations, which may differ in terms of frequency of PALB2 pathogenic mutations, BC incidence rate, and genetic burden of BC. This was necessitated by the insufficient number of eligible studies available for each ancestry or population as seen in Table S1. Moreover, even though some studies included data from multiple ancestries or populations, they reported only overall risk estimates. As more risk estimates become available for a specific ancestry, a separate meta-analysis for that ancestry will likely produce more accurate estimates. In absence of meta-analysis based penetrance estimates of BC for PALB2 mutations, genetic counselling of PALB2 carriers with a family history of breast cancer is challenging (Wesoła and Jeleń, 2017). Counsellors would need to navigate the wide variability of estimates implied by the curves in Figure 2. The consensus estimate reported here is a principled compromise, whose credible band is far less wide than the variability of the individual risk estimates that were used to obtain it, owing to the strength of meta-analytic methodology. Thus it can help clinicians streamline the process of counselling carriers of pathogenic PALB2 mutations to mitigate their BC risk. Our final age-specific meta-analytic estimates have been incorporated into the PanelPRO R package — a Mendelian risk prediction model which estimates carrier probabilities and cancer risk for individuals with family history by integrating a large number of gene mutations and a wide range of potential cancer types based on available allele frequencies and penetrances (Lee et al., 2021; Ruberu et al., 2024).

## Data Availability

All data produced in the present study are available on Table 1 of the manuscript.

## 5 Acknowledgment

This work is supported by NIH grant R03CA242562-01A1. We are thankful to Dr. Kevin Hughes and Dr. Kanhua Yin for their help in identifying relevant studies for meta-analysis.

## References

Ahearn, T. U., Pal Choudhury, P., Derkach, A., Wiafe-Addai, B., Awuah, B., Yarney, J., et al. (2022). Breast cancer risk in women from Ghana carrying rare germline pathogenic mutations. Cancer Epidemiology, Biomarkers & Prevention, 31(8):1593–1601.

Aloraifi, F., McCartan, D., McDevitt, T., Green, A. J., Bracken, A., and Geraghty, J. (2015). Protein-truncating variants in moderate-risk breast cancer susceptibility genes: A meta-analysis of high-risk case-control screening studies. Cancer Genetics, 208(9):455–463.

Antoniou, A. C., Casadei, S., Heikkinen, T., Barrowdale, D., Pylkäs, K., Roberts, J., et al. (2014). Breast-cancer risk in families with mutations in PALB2. The New England Journal of Medicine, 371(6):497–506.

Bogdanova, N., Sokolenko, A. P., Iyevleva, A. G., Abysheva, S. N., Blaut, M., Bremer, M., et al. (2011). PALB2 mutations in German and Russian patients with bilateral breast cancer. Breast Cancer Research and Treatment, 126(2):545–550.

Casadei, S., Norquist, B. M., Walsh, T., Stray, S., Mandell, J. B., Lee, M. K., et al. (2011). Contribution of inherited mutations in the BRCA2-interacting protein PALB2 to familial breast cancer. Cancer Research, 71(6):2222–2229.

Cybulski, C., Kluzńiak, W., Huzarski, T., Wokołorczyk, D., Kashyap, A., Jakubowska, A., et al. (2015). Clinical outcomes in women with breast cancer and a PALB2 mutation: A prospective cohort analysis. The Lancet Oncology, 16(6):638–644.

Dansonka-Mieszkowska, A., Kluska, A., Moes, J., Dabrowska, M., Nowakowska, D., Niwinska, A., et al. (2010). A novel germline PALB2 deletion in Polish breast and ovarian cancer patients. BMC Medical Genetics, 11(1):1–9.

Deng, Z., Yin, K., Bao, Y., Armengol, V. D., Wang, C., Tiwari, A., et al. (2019). Validation of a semiautomated natural language processing–based procedure for meta-analysis of cancer susceptibility gene penetrance. JCO Clinical Cancer Informatics, 3:1–9.

Díaz-Zabala, H., Guo, X., Ping, J., Wen, W., Shu, X.-O., Long, J., et al. (2022). Evaluating breast cancer predisposition genes in women of african ancestry. Genetics in Medicine, 24(7):1468– 1475.

Dorling, L., Carvalho, S., Allen, J., González-Neira, A., Luccarini, C., WahlstrÖm, C., et al. (2021). Breast cancer risk genes-association analysis in more than 113,000 women. The New England Journal of Medicine, 384(5):428–439.

Erkko, H., Dowty, J. G., Nikkilá, J., Syrjákoski, K., Mannermaa, A., Pylkäs, K., et al. (2008). Penetrance analysis of the PALB2 c.1592delT founder mutation. Clinical Cancer Research, 14(14):4667–4671.

Felix, G. E., Guindalini, R. S. C., Zheng, Y., Walsh, T., Sveen, E., Lopes, T. M. M., et al. (2022). Mutational spectrum of breast cancer susceptibility genes among women ascertained in a cancer risk clinic in Northeast Brazil. Breast Cancer Research and Treatment, 193(2):485–494.

Heikkinen, T., Kärkkäinen, H., Aaltonen, K., Milne, R. L., Heikkilä, P., Aittomäki, K., et al. (2009). The breast cancer susceptibility mutation PALB2 1592delT is associated with an aggressive tumor phenotype. Clinical Cancer Research, 15(9):3214–3222.

Hofstatter, E. W., Domchek, S. M., Miron, A., Garber, J., Wang, M., Componeschi, K., et al. (2011). PALB2 mutations in familial breast and pancreatic cancer. Familial Cancer, 10(2):225–231.

Jones, S., Hruban, R. H., Kamiyama, M., Borges, M., Zhang, X., Parsons, D. W., et al. (2009). Exomic sequencing identifies PALB2 as a pancreatic cancer susceptibility gene. Science, 324(5924):217–217.

Kraft, P. and Thomas, D. C. (2000). Bias and efficiency in family-based gene-characterization studies: Conditional, prospective, retrospective, and joint likelihoods. The American Journal of Human Genetics, 66(3):1119–1131.

Kurian, A. W., Hughes, E., Handorf, E. A., Gutin, A., Allen, B., Hartman, A.-R., et al. (2017). Breast and ovarian cancer penetrance estimates derived from germline multiple-gene sequencing results in women. JCO Precision Oncology, 1:1–12.

Landrum, M., Lee, J., Benson, M., Brown, G., Chao, C., Chitipiralla, S., et al. (2018). Clinvar: improving access to variant interpretations and supporting evidence. Nucleic Acids Research, 46(D1):D1062–D1067.

Lee, G., Liang, J. W., Zhang, Q., Huang, T., Choirat, C., Parmigiani, G., et al. (2021). Multi-syndrome, multi-gene risk modeling for individuals with a family history of cancer with the novel R package PanelPRO. eLife, 10:e68699.

Marabelli, M., Cheng, S. C., and Parmigiani, G. (2016). Penetrance of ATM gene mutations in breast cancer: A meta-analysis of different measures of risk. Genetic Epidemiology, 40(5):425– 431.

Momozawa, Y., Iwasaki, Y., Parsons, M. T., Kamatani, Y., Takahashi, A., Tamura, C., et al. (2018). Germline pathogenic variants of 11 breast cancer genes in 7,051 Japanese patients and 11,241 controls. Nature Communications, 9(1):1–7.

Plichta, J. K., Griffin, M., Thakuria, J., and Hughes, K. S. (2016). What’s new in genetic testing for cancer susceptibility? Oncology (Williston Park, N.Y.), 30(9):787–799.

Poumpouridou, N. and Kroupis, C. (2012). Hereditary breast cancer: beyond BRCA genetic analysis; PALB2 emerges. Clinical Chemistry and Laboratory Medicine, 50(3):423–434.

R Core Team (2020). R: A Language and Environment for Statistical Computing. R Foundation for Statistical Computing, Vienna, Austria.

Rahman, N., Seal, S., Thompson, D., Kelly, P., Renwick, A., Elliott, A., et al. (2007). PALB2, which encodes a BRCA2-interacting protein, is a breast cancer susceptibility gene. Nature Genetics, 39(2):165–167.

Ruberu, T. L. M., Braun, D., Parmigiani, G., and Biswas, S. (2024). Bayesian meta-analysis of penetrance for cancer risk. Biometrics. Accepted; discussants invited by the journal. 2304.01912 [STAT.ME].

Tung, N., Battelli, C., Allen, B., Kaldate, R., Bhatnagar, S., Bowles, K., et al. (2015). Frequency of mutations in individuals with breast cancer referred for BRCA1 and BRCA2 testing using next-generation sequencing with a 25-gene panel. Cancer, 121(1):25–33.

Wesoła, M. and Jeleń, M. (2017). The risk of breast cancer due to PALB2 gene mutations. Advances in Clinical and Experimental Medicine, 26(2):339–342.

Zheng, Y., Walsh, T., Gulsuner, S., Casadei, S., Lee, M. K., Ogundiran, T. O., et al. (2018). Inherited breast cancer in Nigerian women. Journal of Clinical Oncology, 36(28):2820–2825.

