## Supplement for "Meta-Analysis of Breast Cancer Risk for Individuals with PALB2 Pathogenic Variants"

### Summary of the Meta-Analysis Method

Our meta-analysis approach proposed in Ruberu et al. (2024) can integrate different types of risk measures such as age-specific penetrances, OR, RR, and SIR accounting for uncertainties involved in such a synthesis. Specifically, we assume that, irrespective of the reporting modality, the cumulative penetrance $F_s(t|\kappa_s, \lambda_s)$ at age $t$ for study $s$ is given by the c.d.f. of a Weibull distribution with shape parameter $\kappa_s$ and scale parameter $\lambda_s$. For each modality, we assume that the reported risk estimate follows a normal distribution whose mean is expressed in terms of Weibull distribution (and hence its parameters $\kappa_s$ and $\lambda_s$) and variance is fixed at what was reported by the study. This enables expressing the likelihood for each study in terms of penetrance parameters as in Marabelli et al. (2016). Furthermore we assume that studies have no patient overlap and are conditionally independent given the study-specific parameters specified below. Our full likelihood is then simply the product of likelihoods from each study.

We employed a Bayesian hierarchical model where the $\kappa_s$ and $\lambda_s$ parameters are assumed to follow gamma priors with their parameters having uniform hyper-priors each with pre-specified lower and upper limits $(l, u)$. Specifically, we used the following hierarchical priors: $\kappa_s \sim \text{Gamma}(a, b)$, $\lambda_s \sim \text{Gamma}(c, d)$, where $a$ and $c$ are shape parameters, $b$ and $d$ are scale parameters, and $a \sim \text{U}(7.5, 27.5)$, $b \sim \text{U}(0.15, 0.25)$, $c \sim \text{U}(43, 63)$, and $d \sim \text{U}(1.32, 2.02)$. To choose the limits of the uniform distributions, our main consideration was that the penetrance values (from Weibull distributions) at ages 20-80 resulting from those choices ensured a wide coverage while avoiding unrealistic values. For instance, a penetrance value of 90% at age 20 is deemed unrealistic.

We started by fixing the limits $(l, u)$ for each hyper-parameter, generated 50 values for $a$, $b$, $c$, and $d$, then using those values simulated 50 $\kappa_s$ and $\lambda_s$, and computed penetrance values from resulting Weibull($\kappa_s$, $\lambda_s$) distributions. We repeated this process by varying the limits and chose the ones that best meet our criteria by drawing histograms of the resulting distributions at ages 20, 40, 60, and 80.

To implement the method we used a Markov chain Monte Carlo (MCMC) algorithm. After estimating the posterior distributions of all parameters, we estimated the final meta-analysis penetrance curve. Specifically, for MCMC iteration $t$: (1) We computed $\kappa^{(t)} = a^{(t)} * b^{(t)}$ and $\lambda^{(t)} = c^{(t)} * d^{(t)}$ and (2) Used the Weibull($\kappa^{(t)}, \lambda^{(t)}$) cdf at ages 40, 50, 60, 70, and 80 as penetrance estimates at the $t^{th}$ iteration. Finally, for each age, the mean of the penetrance values over all iterations are computed, which serves as the meta-analysis estimate. We also obtained credible intervals (CrI) using these posterior distributions of penetrances at each age.

We also investigated how sensitive our results are to the choice of fixed hyper-parameters. We explored two other sets of hyper-priors — one set allows for a wider range of penetrance values at each age compared to our chosen hyper-parameters as described above while the other would somewhat restrict the range of penetrance values. Both sets still ensure that the resulting penetrance values are in realistic ranges. The results of the sensitivity analysis showed that there were no substantial differences among penetrance values generated by different sets of hyper parameters. Please refer Ruberu et al. (2024) for a detailed description of the method.

Table S1: Details about studies included in the meta-analysis of BC penetrance for PALB2 mutations.

| Index | Study | Country | Ancestry | | # cases | # controls | Mutation Prevalence |
|---|---|---|---|---|---|---|---|
| 1 | Antoniou et al. | Multiple[a] | Not Reported | | 229 | 82 | NA[c] |
| 2 | Erkko et al. | Finland | Not Reported | | 90 | 123 | NA[c] |
| 3 | Rahman et al. | UK | Non UK ethnic groups<br>were removed from cases.<br>controls were 97% White | | 923 | 1084 | 0.005 |
| 4 | Kurian et al. | USA | Western/Northern European<br>Central/Eastern European<br>Latin American/Caribbean<br>African<br>Native American<br>Asian<br>Ashkenazi<br>Near/Middle Eastern | 57%<br>14%<br>9%<br>9%<br>4%<br>3%<br>2%<br>1% | 26384 | 69177 | 0.005 |
| 5 | Momozawa et al. | Japan | Japanese ancestry | | 7051 | 11241 | 0.002 |
| 6 | Díaz-Zabala et al. | USA | African American | | 1117 | 2169 | 0.004 |
| 7 | Cybulski et al. | Poland | Not Reported | | 12529 | 4702 | 0.007 |
| 8 | Dorling et al. | Multiple[b] | European<br>Asian | | 47522 | 50475 | 0.003 |
| 9 | Heikkinen et al. | Finland | Not Reported | | 1274 | 1079 | 0.004 |
| 10 | Ahearn et al. | Ghana | African | | 871 | 1563 | 0.004 |
| 11 | Zheng et al. | Nigeria | African | | 1136 | 997 | 0.005 |
| 12 | Felix et al. | Brazil | White<br>African-descended<br>Other | 19%<br>77%<br>4% | 171 | 119 | 0.010 |

[a] Families from 14 participating research centers across the world

[b] Based on 30 studies in breast cancer association consortium (BCAC) unselected for family history.

[c] Penetrance computation only use carriers
